# Hemoglobin concentration maintained despite reduced erythrocyte count and hematocrit during exercise under hyperbaric oxygen conditions in healthy adult males: A randomized crossover trial

**DOI:** 10.1101/2025.09.29.25336665

**Authors:** Takehira Nakao, Atsushi Saito, Takahiro Adachi, Jun Fukuda, Tadanori Fukada, Kaori Iino-Ohori, Kensuke Iwasa, Miki Igarashi, Keisuke Yoshikawa

**Author notes:** **Corresponding author: Takehira Nakao or Keisuke Yoshikawa** Takehira Nakao, M. Sc. Faculty of Human Science, Kyushu Sangyo University, 3-1 Matsukadai 2, Higashi-ku, Fukuoka, 813-0004, Japan Keisuke Yoshikawa, Ph.D. Department of Pharmacology, Faculty of Medicine, Saitama Medical University, 38 Moro-hongo, Moroyama-machi, Iruma-gun, Saitama 350-0495, Japan.

## Abstract

Hyperbaric oxygen (HBO), physical exercise, and eicosapentaenoic acid (EPA) intake combined may have beneficial effects on blood components and vascular endothelial function. To test this hypothesis, we conducted a randomized crossover trial involving healthy adult male participants. Participants were assigned to three groups: two performed exercise under HBO or normobaric normoxia (NN) conditions while taking EPA (HBO + Ex + EPA and NN + Ex + EPA), and one received EPA without exercise (EPA control). Each exercise-based intervention lasted for 4 weeks, with a 1-week washout period before crossover into the other environment. Blood parameters and endothelial function were measured before and after the interventions. In the HBO + Ex + EPA group, hemoglobin (Hb) levels remained unchanged despite reductions in the red blood cell (RBC) count and hematocrit (Ht). No significant changes were observed in the RBC or Ht in the NN + Ex + EPA or EPA control groups, suggesting that the reductions were likely attributable to exercise performed under HBO conditions. No significant changes were observed in other blood parameters (blood sugar, glutamate oxaloacetate transaminase, glutamate pyruvate transaminase, and gamma-glutamyl transpeptidase concentrations), blood lipid concentrations (triglycerides and high- and low-density lipoprotein cholesterol), or indicators of vascular endothelial function (baseline brachial artery diameter, maximum brachial artery diameter, flow-mediated dilation percentage, and vascular wall shear rate) in either of the exercise groups. In the EPA control group, triglyceride levels significantly decreased and high-density lipoprotein cholesterol levels increased, whereas no significant changes were observed in other blood markers or indicators of vascular endothelial function. In the current study, Hb levels were maintained despite reductions in the RBC count and Ht during exercise under HBO conditions. This study provides valuable fundamental insights into the effects of exercise performed in extreme physiological environments, such as HBO conditions, on blood homeostasis and vascular function.

## Introduction

A hyperbaric oxygen (HBO) environment, physical exercise, and intake of n-3 polyunsaturated fatty acids (PUFAs) may have beneficial effects on blood components and vascular endothelial function. Vascular endothelial cells play important roles in the maintenance of vascular homeostasis, regulation of blood circulation, and secretion of bioactive compounds such as nitric oxide (NO), endothelin-1, and interleukins (ILs) (1, 2). The cells also secrete vascular endothelial growth factor, which promotes angiogenesis, produces antioxidants, and removes reactive oxygen species (ROS) via the enzyme catalase (3). The impairment of vascular endothelial function not only increases the risk of lifestyle-related diseases such as hypertension, cerebrovascular disease, and dyslipidemia, but also contributes to motor dysfunction and the progression of dementia (4, 5). Therefore, the establishment of physiological and nutritional approaches for the maintenance and improvement of endothelial function is an important issue in the fields of sports science and medicine.

Physical exercise in a normal environment promotes angiogenesis and improves vascular repair by increasing the recruitment of endothelial progenitor cells (6). Aerobic exercise reportedly increases NO production in the vascular endothelium and promotes vasodilation (7). Exercise also has a considerable significant effect on blood components, reducing the expression of pro-inflammatory cytokines such as tumor necrosis factor-α and IL-6, and reportedly contributes to the suppression of inflammatory responses (8). Research on HBO environments has mostly been conducted using animal models such as rats, sheep, and rabbits (9, 10), and few studies involving humans have been conducted. The combination of an HBO environment and high-intensity interval training moderately induce ROS; promotes NO production, which results in improved vascular endothelial function (11). However, comprehensive investigations into the synergistic effects of HBO environments and exercise are limited, particularly with respect to healthy young adults. In addition, the effects of exercise in an HBO environment on blood components remain unclear. Furthermore, the long-term HBO adaptation mechanisms and optimal exercise protocols to enhance vascular endothelial function are yet to be established.

The intake of n-3 PUFAs may improve vascular function through anti-inflammatory effects and improved blood flow (12, 13). Their intake increases serum concentrations of free n-3 PUFAs, improves microvascular endothelium-dependent vasodilation via the reduction of pro-inflammatory cytokines and increase of anti-inflammatory cytokines (14), and significantly increases flow-mediated dilation (FMD) (15). In particular, eicosapentaenoic acid (EPA) has been suggested to have anti-inflammatory and anti-thrombotic effects, as well as reducing platelet aggregation (4, 5).

Therefore, the combination of appropriate physical exercise and EPA intake in an HBO environment may have a beneficial synergistic effect in terms of improvements in vascular endothelial function and blood components. In this study, we aimed to comprehensively investigate those potential effects.

## Materials and Methods

### Study design

This study was conducted as a randomized crossover trial (Fig.1). Participants were divided into three groups. Two of the groups performed exercise stress tests under HBO or normobaric normoxia (NN) conditions while taking EPA daily, (HBO + Ex + EPA) and (NN + Ex + EPA) groups, respectively. Subsequently, the groups switched environments and repeated the tests. The third group took EPA intake but not engage in exercise, as EPA control group in NN environment. Each test period was 4 weeks, followed by a 1-week washout period, based on previous studies on the half-life of the plasma EPA concentration (16). Blood samples were collected and various measurements were performed before, during, and after the experiments. The study was conducted in accordance with the tenets of the Declaration of Helsinki, approved by the Ethics Committee of Kyushu Sangyo University (approval number: 2023-0006), and registered in the University Hospital Medical Information Network (ID: UMIN000057211). Written informed consent was obtained from all participants.

**Figure 1.**
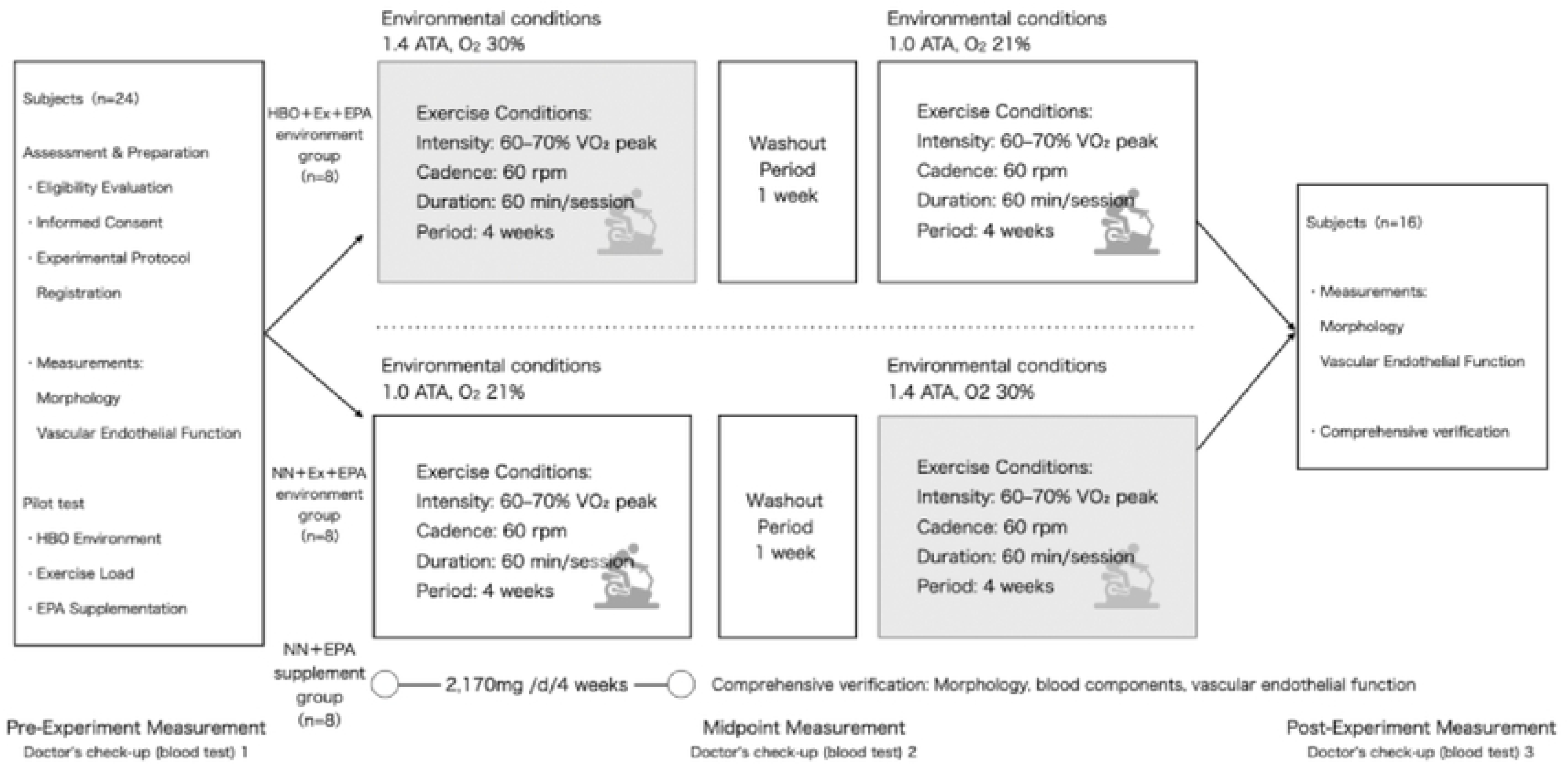
Experimental design (randomized crossover trial)

### Participants

Participants for this study were recruited during the period from April 25 to June 18, 2024. We recruited 24 healthy adult males (mean ± SD age: 20.9 ± 1.4 years, height: 171.4 ± 5.6 cm, weight: 64.1 ± 11.6 kg, body mass index [BMI]: 21.8 ± 3.6 kg/m^2^) through public recruitment, randomly dividing them into HBO + Ex + EPA, NN + Ex + EPA, and EPA control groups (n = 8 each). The inclusion criteria were as follows: (1) no history of cardiovascular or metabolic disease, (2) no history of smoking, (3) the ability to perform strenuous exercise and take of EPA-containing supplements during the study period, and (4) ability to adapt to a hyperbaric environment. The exclusion criteria were as follows: (1) age < 20 years, (2) excessive exercise habits, and (3) intake of specific foods and/or supplements. Participants were instructed to avoid substantial changes in their diet and lifestyle throughout the study period.

### Experimental environment

#### 1) Environments

For the HBO environment, we maintained an absolute pressure of 1.41 ± 0.008 ATA, an oxygen concentration of 29–31 %, a temperature of 21.7 ± 0.7 °C, and a humidity of 73.5 ± 4.6 %. For the NN environment, we maintained an absolute pressure of 1.00 ± 0.998 ATA, an oxygen concentration of 20.9 %, a temperature of 22.1 ± 1.1 °C, and a humidity of 70.5 ± 6.5 %. The HBO environment was created in an artificial environmental control room (Japan Barometric Bulk Industry Co., Ltd., Shizuoka, Japan), with pressurization and depressurization performed at a rate of 0.07 ATA/min. A bicycle ergometer (STB-3400, Nihon Kohden Co., Ltd., Tokyo, Japan) was installed in the room. The NN environment was established in a standard laboratory with similar equipment.

#### 2) Suitability assessment and pre-measurement

The participants underwent a preliminary interview, an exercise stress test (with measurement of the peak oxygen uptake [VO_2peak_]), and an assessment of their ability to adapt to a hyperbaric environment before being enrolled in the experimental protocol.

#### 3) Medical examination, blood sampling, and blood component measurements

A medical examination was conducted by a physician, and blood samples were collected at three time points: the early, middle, and late phases of the experiment. Venous blood was drawn using butterfly needles equipped with needlestick prevention devices (NIPRO Co., Ltd, Settsu, Japan) and vacuum blood collection tubes (EDTA-2Na, 5 mL; TERUMO Co., Ltd., Tokyo, Japan). The collected samples were stored in a refrigerator at 5 °C for approximately two hours and subsequently analyzed by BML Inc. (Tokyo, Japan). The following blood components were measured: blood sugar (BS), red blood cells (RBCs), hemoglobin (Hb), hematocrit (Ht), glutamate oxaloacetate transaminase (GOT), glutamate pyruvate transaminase (GPT), gamma-glutamyl transpeptidase (γ-GTP), triglycerides (TGs), high-density lipoprotein (HDL) cholesterol, and low-density lipoprotein (LDL) cholesterol. The mean corpuscular hb (MCH) and MCH concentration (MCHC) were calculated using following formulas: MCH (pg) = Hb (g/dL) × 10/RBC (10^4^/μL) and MCHC (g/dL) = Hb (g/dL) × 100/Ht (%).

#### 4) Morphometry

Participant’s height was measured using a standard stadiometer (A&D Co. Ltd., Tokyo, Japan). Body weight, body composition parameters (including fat-free mass (FFM), fat mass (FM), and body fat percentage (% fat)), and basal metabolic rate (BMR) were assessed using a dedicated bioelectrical impedance device (InBody 770; InBody Japan Co., Ltd., Tokyo, Japan). Based on these measurements, the BMI, FFM index (FFMI), and FM index (FMI) were calculated using the following formulas: BMI = weight (kg) / height² (m²); FMI = FM (kg) / height² (m²); FFMI = FFM (kg) / height² (m²). Additionally, systolic blood pressure (SBP) and diastolic blood pressure (DBP) were measured using an automated upper-arm blood pressure monitor (HBP-1300; OMRON Co., Ltd., Kyoto, Japan), employing the oscillometric method.

#### 5) Exercise load

The exercise intensity for the bicycle ergometer test was set at 60–70 % of peak oxygen uptake (VO₂_peak_), with a pedaling rate of 60 rpm, a duration of 60 minutes per session, and a frequency of twice per week for four weeks. The VO₂_peak_ was estimated before the intervention via a respiratory metabolic monitoring system (AE-310S; Minato Medical Science Co., Ltd., Osaka, Japan), using breath-by-breath analysis. Exercise under HBO conditions was performed for 60 minutes once the predetermined absolute pressure was reached. The time required for pressurization and decompression was approximately 20 minutes each. Participants in the HBO group exercised under HBO conditions during the first phase and under NN conditions during the second phase, whereas those in the NN group followed the reverse sequence.

#### 6) EPA intake

During the study period, participants were instructed to take 2,170 mg of a highly concentrated EPA supplement (Bizen Chemical Co., Ltd., Akaiwa, Japan) in capsule form daily. The EPA supplementation period overlapped with the exercise period in both environments. Supplementation was discontinued before the start of the study and during the washout period. To ensure compliance and monitor gastrointestinal health, participants were asked to report their supplementation status and stool characteristics online each day throughout the study period.

#### 7) Evaluation of vascular endothelial function

Vascular endothelial function was assessed using FMD via a B-mode ultrasound imaging device (UNEX EF 38G; UNEX Co., Ltd., Nagoya, Japan) equipped with a 7.5 MHz linear array transducer. After the participant rested in the supine position for 5 min, a probe was placed on the brachial artery, at the midpoint between the right acromion and the olecranon, and a longitudinal scan was performed. Following measurement of the baseline brachial artery diameter, the resting heart rate and blood pressure were recorded. A pneumatic cuff was subsequently placed around the participant’s forearm and inflated to approximately 160 mmHg within 10 seconds, and vascular occlusion was maintained for 5 minutes. After release of the occlusion, FMD was performed while the participant remained in the same position for 2 minutes. The following parameters were evaluated: the baseline diameter (BD), maximum diameter (MD), FMD percentage (%FMD), and vascular wall shear rate (SR).

### Statistics

Statistical analyses were performed using SPSS Statistics version 27 (IBM Corp., Armonk, NY). Two-way repeated measures analyses of variance (ANOVA) were used to examine the effects of environmental conditions (HBO vs. NN) and time (before and after the intervention). A preliminary power analysis was conducted for the F-test for the interaction by using GPower (version 3.1), with the significance level set at α = 0.05 and power at 80 %, considering the two environmental conditions and two measurement time points. The results indicated that 24 participants were required to detect an effect size of 0.25 (medium). However, as this study was designed as an early-stage exploratory (pilot) study, although the number of participants did not meet the recommended threshold, a sufficient degree of statistical power was achieved by adopting a randomized crossover design and minimizing individual differences among the participants. Furthermore, to account for the potential lack of statistical power, we did not rely solely on statistical significance (*p* value) to interpret the results, but also reported the effect size (η^2^ and Cohen’s d) and 95 % confidence intervals (CIs) to assess precision and practical significance. As the control group, which was not included in the crossover study, was small (n = 8), and considering the possibility that the data were not normality distributed, the Wilcoxon signed rank test was used to compare parameters before and after the intervention, and the significance (*p* value) and effect size (r value) are shown. In all statistical analyses, the significance level was set at *p* < 0.05 for two-sided tests.

## Results

### Physical characteristics

Table 1 presents the characteristics of participants in the HBO + Ex + EPA and NN + Ex + EPA groups before and after the intervention.

**Table 1.** Morphology, body composition, basal metabolic rate, and blood pressure.

| Measurement |  | HBO + Ex + EPA |  | NN + Ex + EPA |  | *P*-value<br>Group*time |
| --- | --- | --- | --- | --- | --- | --- |
|  |  | Pre | Post | Pre | Post |  |
| Height | cm | 170.4 ± 5.9 | 170.3 ± 5.9 | 170.7 ± 6.0 | 170.4 ± 5.9 | 0.113 |
| Weight | kg | 65.1 ± 11.4 | 65.8 ± 11.8 | 64.5 ± 12.0 | 64.9 ± 11.0 | 0.588 |
| BMI | kg/m<sup>2</sup> | 22.3 ± 2.9 | 22.5 ± 3.1 | 22.0 ± 3.3 | 22.3 ± 2.9 | 0.975 |
| FFM | kg | 52.2 ± 6.9 | 52.7 ± 7.2 | 52.3 ± 7.3 | 52.0 ± 6.5 | 0.108 |
| FM | kg | 12.9 ± 5.3 | 13.1 ± 5.5 | 12.2 ± 5.5 | 13.0 ± 5.5 | 0.260 |
| %Fat | % | 19.2 ± 4.8 | 19.3 ± 4.9 | 18.3 ± 4.8 | 19.4 ± 5.1 | 0.137 |
| FFMI | kg/m<sup>2</sup> | 17.9 ± 1.6 | 18.1 ± 1.6 | 17.9 ± 1.7 | 17.9 ± 1.5 | 0.329 |
| FMI | kg/m<sup>2</sup> | 4.4 ± 1.6 | 4.5 ± 1.7 | 4.2 ± 1.7 | 4.4 ± 1.7 | 0.181 |
| BMR | kcal | 1497.2 ± 149.8 | 1507.5 ± 155.7 | 1499.5 ± 157.2 | 1492.3 ± 140.2 | 0.112 |
| SBP | mmHg | 115.7 ± 9.6 | 117.6 ± 7.6 | 116.4 ± 7.3 | 114.2 ± 9.5 | 0.107 |
| DBP | mmHg | 61.0 ± 5.9 | 61.4 ± 6.8 | 60.3 ± 7.0 | 61.3 ± 6.9 | 0.806 |

Data are reported as means ± standard deviation.

No significant interaction effects were observed between the environmental conditions and time in terms of height, weight, BMI, body composition parameters, BMR, or blood pressure. The EPA control group exhibited no significant changes in physical characteristics (all *p* > 0.05; data not shown).

### Blood components

Table 2 shows the changes in blood components before and after the experiment under HBO + Ex + EPA and NN + Ex + EPA conditions.

**Table 2.** Characteristics of blood components.

| Measurement |  | HBO + Ex + EPA |  |  | NN + Ex + EPA |  |  | P-value |
| --- | --- | --- | --- | --- | --- | --- | --- | --- |
|  |  | Pre |  | Post | Pre |  | Post | Group*time |
| BS | mg/dL | 97.81 ± 10.69 |  | 99.75 ± 8.33 | 99.75 ± 10.23 |  | 100.44 ± 13.35 | 0.819 |
| GOT | U/L | 20.69 ± 5.74 |  | 21.56 ± 7.29 | 20.19 ± 5.55 |  | 21.06 ± 6.10 | 1.000 |
| GPT | U/L | 23.63 ± 12.03 |  | 27.25 ± 18.21 | 22.31 ± 12.40 |  | 24.69 ± 12.65 | 0.773 |
| γ-GTP | U/L | 20.50 ± 6.58 |  | 20.62 ± 7.07 | 19.37 ± 6.29 |  | 20.00 ± 7.14 | 0.704 |
| GOT/GPT |  | 1.00 ± 0.33 |  | 0.95 ± 0.40 | 1.03 ± 0.38 |  | 0.96 ± 0.27 | 0.778 |
| RBC | 10<sup>4</sup>/μL | 520.30 ± 29.36 |  | 506.69 ± 27.49† | 507.00 ± 31.31 |  | 512.81 ± 30.01 | 0.012* |
| Ht | % | 46.40 ± 2.07 |  | 45.28 ± 1.85† | 45.38 ± 2.36 |  | 45.85 ± 2.36 | 0.019* |
| Hb | g/dL | 15.35 ± 0.90 |  | 15.03 ± 0.68 | 15.09 ± 0.92 |  | 15.16 ± 0.92 | 0.057 |
| TG | mg/dL | 94.19 ± 62.45 |  | 91.56 ± 41.0 | 89.75 ± 36.10 |  | 95.69 ± 63.58 | 0.680 |
| HDL | mg/dL | 56.81 ± 12.65 |  | 53.31 ± 10.5 | 53.06 ± 9.09 |  | 55.06 ± 12.94 | 0.028* |
| LDL | mg/dL | 89.81 ± 22.87 |  | 87.12 ± 25.4 | 88.94 ± 25.01 |  | 88.00 ± 30.63 | 0.665 |
| LDL/HDL |  | 1.69 ± 0.75 |  | 1.73 ± 0.7 | 1.74 ± 0.71 |  | 1.73 ± 0.99 | 0.649 |

Data are reported as means ± standard deviation. \*indicates a significant interaction
between group and time from pre- to post- intervention (p < 0.05). †indicates a
significant difference within the group between pre- and post-values (p < 0.05).

No interaction between environment and time was observed for BS, GOT, GPT, or γ-GTP levels or the GOT/GPT ratio (all *p* > 0.05). Significant environment–time interactions were observed for the RBC count (F_1,30_ = 7.161, *p* = 0.012, η^2^ = 0.193) and Ht (F_1,30_ = 6.124, *p* = 0.019, η^2^ = 0.170) but not for the Hb level (*p* = 0.057). Paired t-tests for each group revealed significant decreases in the RBC count (from 520.30 ± 29.36×10^4^ cells/μL to 506.69 ± 27.49×10^4^ cells/μL, t_15_ = 2.447, *p* = 0.027, 95 % CI: - 25.01 to -2.25 × 10^4^ cells/μL, d = 0.612) (Fig. 2A) and Ht (46.40 ± 2.07 % to 45.28 ± 1.85 %, t_15_ = 2.287, *p* = 0.037, 95 % CI: 0.076 to 2.162 %, d = 1.957) (Fig. 2B) in the HBO + Ex + EPA group. However, no changes in the Hb level were observed in the HBO + Ex + EPA group (Fig. 2C). Moreover, no significant changes in the RBC count (t_15_ = -1.247, *p* = 0.232, 95 % CI: -6.94 to 18.56×10^4^ cells/μL, d = -0.312), Ht (t_15_ = - 1.130, *p* = 0.276, 95 % CI: -1.350 to 0.416 %, d = 1.660), or Hb level were observed in the NN + Ex + EPA group (Fig. 2). Although no changes were observed in the MCH level or MCHC in the HBO + Ex + EPA group, both parameters exhibited slight (though not statistically significant) decreases in the NN + Ex + EPA group (Fig.3). No interaction between environment and time was observed for the TG concentration, LDL cholesterol concentration, or LDL/HDL cholesterol ratio (all *p* > 0.05) (Table 2). A significant interaction between environment and time was observed for the HDL cholesterol concentration (F_1,30_ = 5.354, *p* = 0.028, η^2^ = 0.151), but the interaction was not significant upon paired t-testing (t_15_ = 1.824, *p* = 0.088, 95 % CI: -0.591 to 7.591, d = 0.777). In the EPA control group, the TG concentration significantly decreased from 130.9 ± 72.8 mg/dL to 79.4 ± 54.3 mg/dL (*p* < 0.016, r = 0.85), and the HDL cholesterol concentration significantly increased from 53.3 ± 13.8 mg/dL to 58. 4 ± 15.4 mg/dL (*p* < 0.028, r = 0.78), whereas no significant changes were observed in other blood components (data not shown).

**Figure 2.**
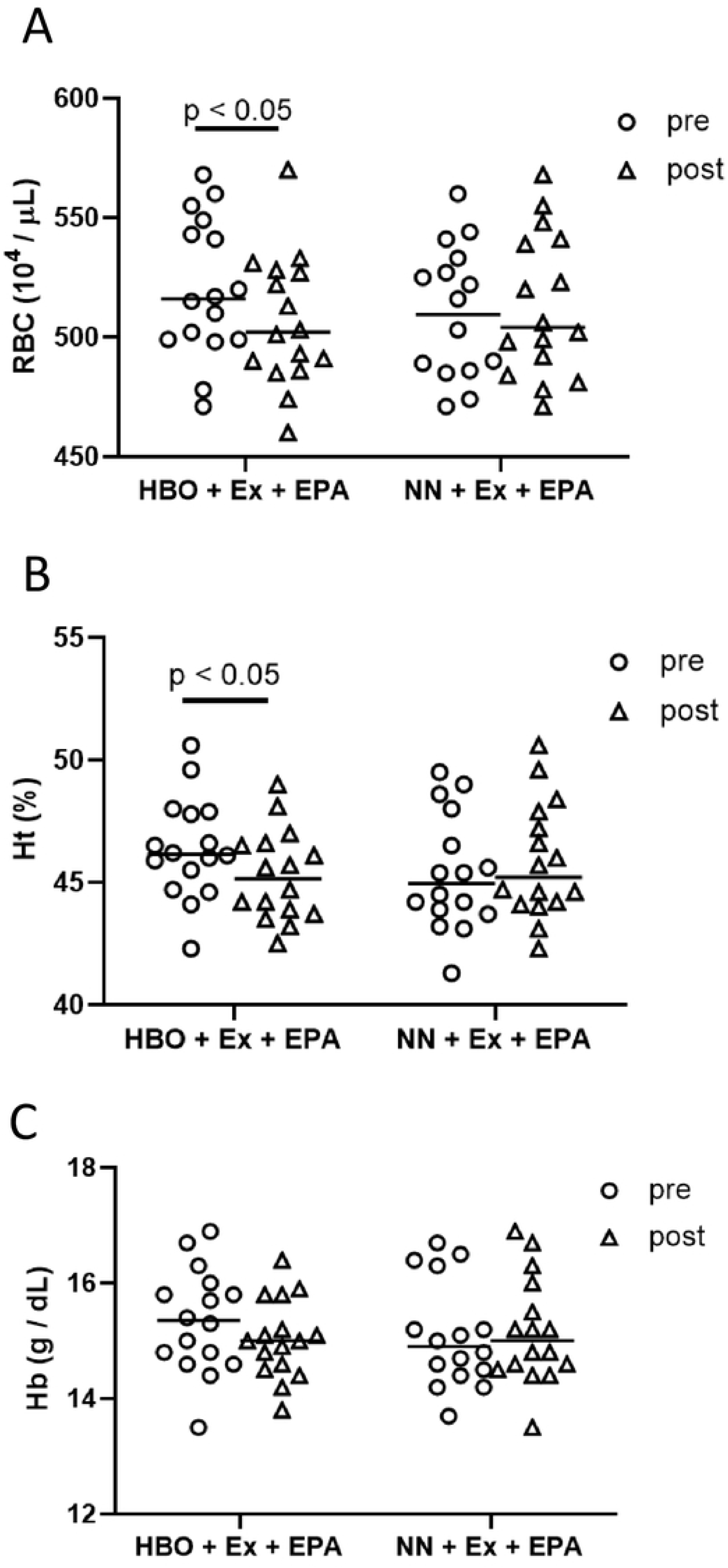
RBC counts (A) and Ht (B) before and after intervention. Statistical analyses were performed using two-way repeated-measures ANOVAs.

**Figure 3.**
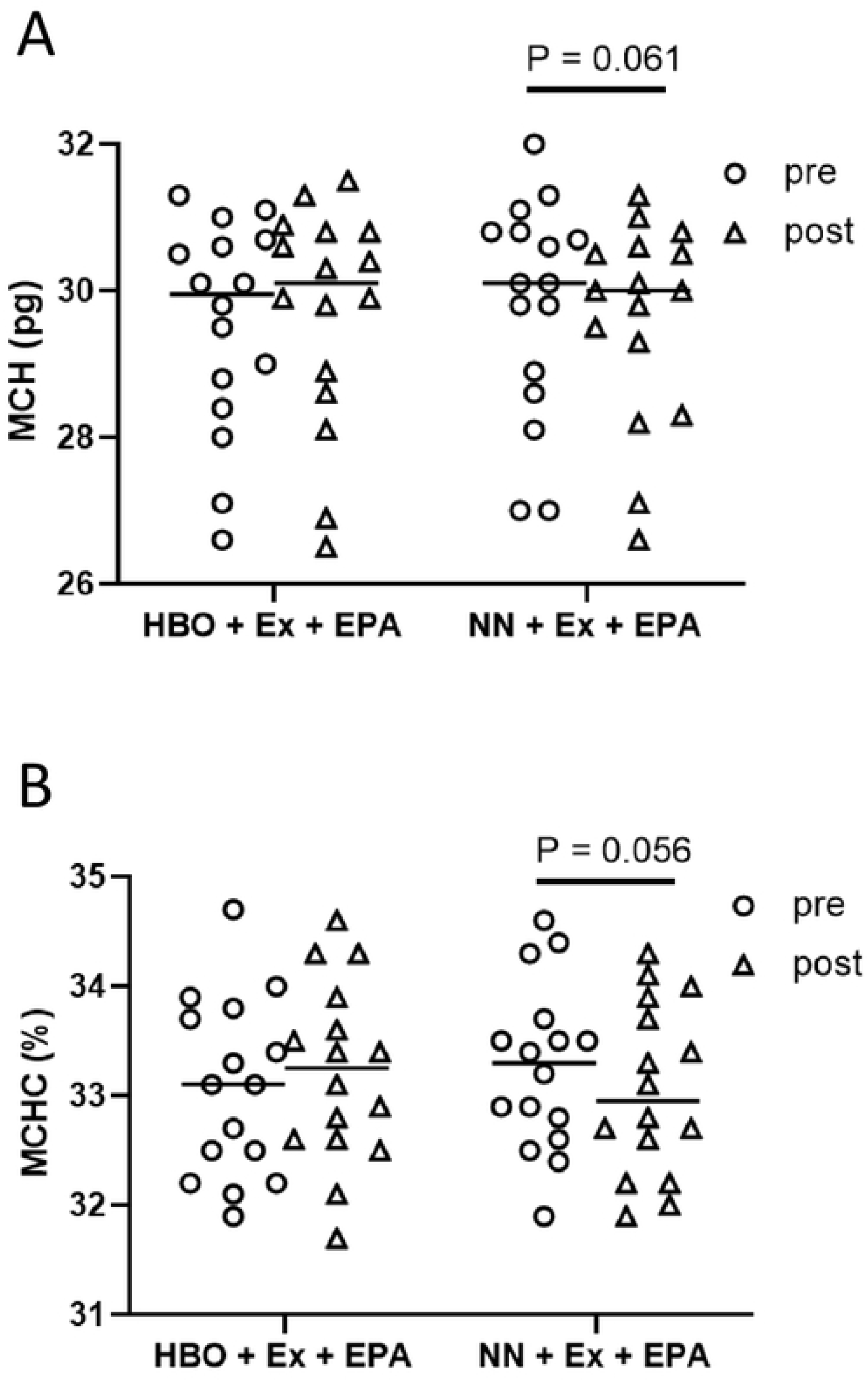
MCH (A) and MCHC (B) before and after the intervention. Statistical analyses were performed using two-way repeated-measures ANOVAs.

### Vascular endothelial function

The arterial vascular functional indices (BD, MD, %FMD, and SR) before and after the experiment in the HBO and NN groups are shown in Table 3.

**Table 3.**
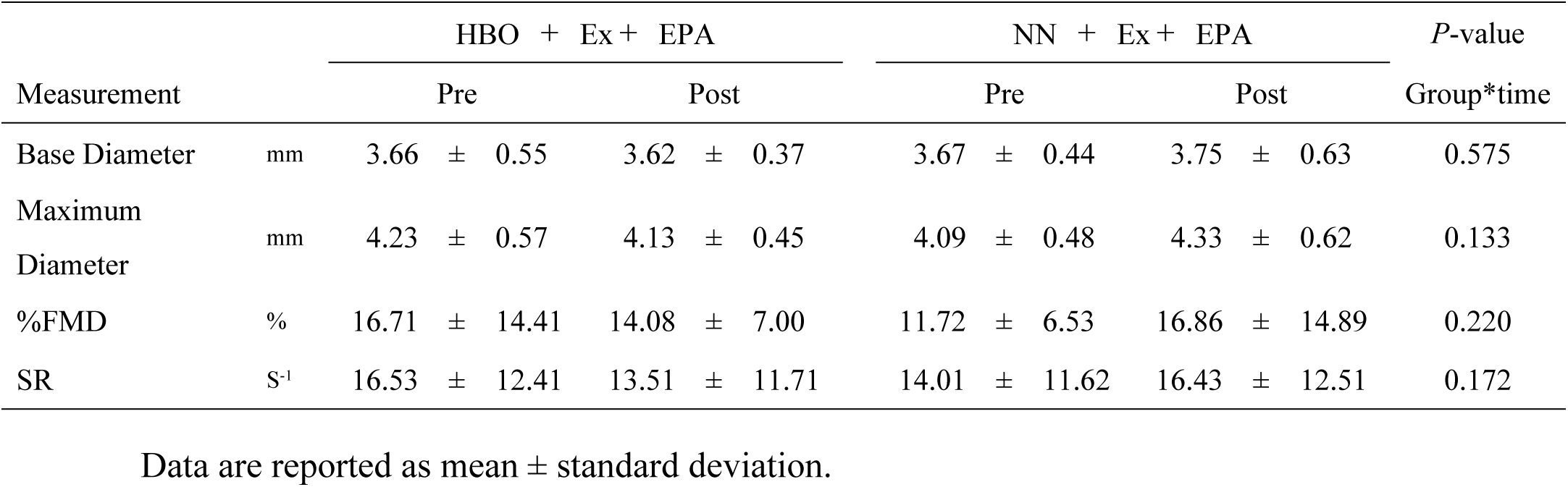
Characteristics of vascular endothelial function.

No interaction between environment and time was observed for BD, MD, %FMD, or SR (*p* > 0.05). Moreover, no statistically significant changes were observed in the EPA control group (*p* > 0.05).

## Discussion

To our knowledge, this is the first comprehensive investigation of the effects of exercise under HBO conditions and EPA supplementation on blood components and vascular endothelial function in humans via a randomized crossover trial. The HBO + Ex + EPA group exhibited a decrease in RBC count and Ht but no change in the Hb level, MCH, or MCHC. In the NN + Ex + EPA group or EPA control group (without exercise), no changes were observed in any of these parameters. Collectively, these results indicate that the reductions in RBC count and Ht were likely attributable to the exercise performed under HBO conditions. Despite the reductions in RBC count and Ht, participants had a maintained Hb levels, MCH, and MCHC by the end of the exercise under HBO conditions. We infer that blood viscosity can be reduced while oxygen-carrying capacity is maintained under such conditions. Although not significantly, the MCH and MCHC seemed to decrease rather than increase in NN + Ex + EPA group. A decrease in the TG concentrations and an increase in the HDL concentration were observed in the EPA control group. No significant changes were observed in other blood parameters (BS, GOT, GPT, or γ-GTP concentrations), blood lipids (TG, HDL cholesterol, or LDL cholesterol concentrations), or vascular endothelial functional indicators (BD, MD, FMD, or SR) under either the exercise groups.

Our results led us to hypothesize that the number of Hb molecules per RBC increased in the HBO exercise; however, the indices of RBC Hb content and concentration (MCH or MCHC, respectively) did not significantly change. These results may be explained by several possible mechanisms. The MCH and MCHC seemed to decrease in the NN + Ex + EPA group. This apparent decrease might have been offset under HBO conditions by a relative increase in Hb content per RBC. A decrease in the RBC count under HBO was previously reported: the increase in arterial oxygen content under HBO conditions suppresses erythropoietin production in the kidneys, leading to a transient decrease in erythropoiesis (17). Additionally, HBO exposure shortens the lifespan of RBCs in rats, resulting in a reduction in the number of senescent RBCs (18). Enhanced mobilization of hematopoietic stem cells in the bone marrow may compensate for the reduction in RBCs by elevating Hb content.

In our study, the EPA control group exhibited a decreased in TG level and an increased HDL cholesterol level, which is consistent with the results of previous studies (19, 20). Although the amount of EPA administered in this study was sufficient to affect lipid metabolism, it had no significantly affect other blood parameters or endothelial function.

Although we investigated the effects of the interventions on vascular endothelial function, no significant changes were observed in either %FMD or SR under any of the experimental conditions. Previous studies have demonstrated that EPA can improve endothelial function via its antioxidant and anti-inflammatory properties (21). Furthermore, Paola Peña-de-la-Sancha (22) recently reported a significant improvement in FMD following EPA and docosahexaenoic acid supplementation in middle-aged men with hypertriglyceridemia. In contrast, we did not observe any beneficial effects of EPA intake or exercise on endothelial function. Several factors may account for this discrepancy. First, under HBO conditions, the rapid increase in the partial pressure of oxygen might have led to the excessive production of ROS, potentially offsetting the antioxidant effects of EPA and exercise (23). Second, as the participants were healthy young men with high baseline endothelial function, the potential for further improvements might have been low. EPA supplementation at a dose of 2,170 mg/day for 8 weeks might not have been sufficient to induce detectable changes in macro-level indices such as FMD and the SR. Furthermore, our HBO environment and exercise conditions might have imposed only mild environmental physiological stress, potentially insufficient to elicit a clear effect on endothelial function. Taken together, these factors likely contributed to the difficulty in detecting significant intervention effects in the present study.

This study holds considerable academic value as it is, to our knowledge, the first randomized crossover trail conducted to comprehensively investigate, in humans, the effects of three factors—an HBO environment, physical exercise, and EPA intake—on blood components and vascular endothelial function. Furthermore, our results have potential applications in the fields of clinical medicine and health and sports sciences. However, this study had several limitations. First, the sample was small, which might have limited the study’s statistical power to detect significant effects. Second, the study was subject to selection bias. All participants were healthy young males, who might have had limited room for improvement in vascular endothelial function. Therefore, caution is required when these results are generalized to other age groups or females. Third, oxidative stress markers were not measured in this study. Because the potential overproduction of ROS in the HBO environment was not directly assessed, further investigations using specific biomarkers are required for a more thorough evaluation of vascular endothelial function. Fourth, further comprehensive and long-term studies are needed to investigate the combined effects of environmental HBO conditions, EPA intake (dosage and frequency), and exercise parameters such as frequency, intensity, duration, and type. In this was a pilot study, we selected mild environmental conditions with an emphasis on safety. This might have limited the physiological effects. In future, researchers should build upon these results by examining the effects of different environmental conditions; measuring oxidative stress markers (e.g., ROS and antioxidant enzymes); expand the study population to include older adults, individuals with pre-existing conditions, and females; and optimizing EPA dosage and exercise parameters.

In conclusion, we investigated the effects of three factors—an HBO environment, physical exercise, and EPA intake—on blood components and vascular endothelial function. We discovered that exercise under HBO conditions significantly decreased the RBC count and Ht but maintained the Hb levels, MCH, and MCHC, suggesting that blood viscosity was reduced without the loss of oxygen-carrying capacity.

In space, microgravity reportedly leads to a reduction in the RBC count (24, 25). The present results may be valuable as fundamental research to understand the maintenance of physical fitness, regulation of hematological homeostasis, and control of oxidative stress in astronauts. In the future, these results may be applied to the management of blood components and the development of training protocols in space medicine.

## Abbreviations

(ATA): atmosphere absolute
(BMR): basal metabolic rate
(BD): baseline diameter
(BMI): body mass index
(DBP): diastolic blood pressure
(EPA): eicosapentaenoic acid
(Ex): exercise
(FM): fat mass
(FMI): fat mass index
(FFM): fat-free mass
(FFMI): fat-free mass index
(FMD): flow-mediated dilation
(%FMD): flow-mediated dilation percentage
(γ-GTP): gamma-glutamyl transpeptidase
(GOT): glutamate oxaloacetate transaminase
(GPT): glutamate pyruvate transaminase
(Ht): hematocrit
(Hb): hemoglobin
(HDL): high-density lipoprotein
(HBO): hyperbaric oxygen
(IL): interleukin
(IL-6): interleukin-6
(LDL): low-density lipoprotein
(MD): maximum diameter
(MCH): mean corpuscular hemoglobin
(MCHC): mean corpuscular hemoglobin concentration
(NO): nitric oxide
(NN): normobaric normoxia
(PUFA): polyunsaturated fatty acids
(ROS): reactive oxygen species
(RBC): red blood cell
(SR): shear rate
(SBP): systolic blood pressure
(TG): triglycerides

## Acknowledgements

The authors sincerely thank all the participants from Kyushu Sangyo University for their valuable cooperation. We also acknowledge Bizen Chemical Co., Ltd. for generously providing EPA supplements.

## Statements & Declarations

### Data availability

The data sets used and analyzed during the current study are available from the corresponding author on reasonable request.

## Author contributions

**Takehira Nakao**: Writing – review & editing, Writing – original draft, Methodology, Investigation, Visualization, Data curation, Funding acquisition, Conceptualization. **Atsushi Saito**: review & editing, Methodology, Data curation, Funding acquisition, Conceptualization, Supervision. **Takahiro Adachi**: Methodology, Investigation, Resources. **Jun Fukuda**: Methodology, Investigation, Resources. **Tadanori Fukada**: Methodology, Investigation, Resources. **Kaori Iino-Ohori**: Writing – review & editing**. Kensuke Iwasa**: Writing – review & editing, Funding acquisition. **Miki Igarashi:** Writing – review & editing, Methodology, Conceptualization. **Keisuke Yoshikawa**: Writing –review & editing, Writing – original draft, Supervision, Funding acquisition, Conceptualization.

## Ethics approval

The study was approved by the Ethics Committee of Kyushu Sangyo University (approval number: 2023-0006).

